# Mapping the Proteomic Landscape of Aortic Aneurysm and Dissection in the Context of Hypertension

**DOI:** 10.64898/2026.01.21.26344570

**Authors:** Jiaqi Hou, Longjiang Wu, Lihua Lin, Meichen Pan, Jing Huang, Jianghua Du, Shujuan Wang, Xingjie Hao, Chen Chen, Qian Liu

**Affiliations:** Department of Forensic Medicine, Tongji Medical College of Huazhong University of Science and Technology, Wuhan, China; Division of Cardiology, Tongji Hospital, Tongji Medical College and State Key Laboratory for Diagnosis and Treatment of Severe Zoonotic Infectious Diseases, Huazhong University of Science and Technology, Wuhan, China; Hubei Key Laboratory of Genetics and Molecular Mechanisms of Cardiological Disorders, Wuhan, China; Department of Epidemiology and Biostatistics, Ministry of Education Key Laboratory of Environment and Health, School of Public Health, Tongji Medical College, Huazhong University of Science and Technology, Wuhan, China

## Abstract

**Background:** Aortic aneurysm and dissection (AAD) are highly lethal conditions for which hypertension serves as a primary risk factor. The limited efficacy of conventional antihypertensive treatments suggests an inadequate understanding of the molecular mechanisms that connect these conditions. Despite their pivotal role in regulating biological functions, the specific proteomic signatures associated with both hypertension and AAD have not been extensively investigated. This study sought to conduct a comprehensive mapping of the plasma proteome to identify novel biomarkers and therapeutic targets for AAD within a hypertensive cohort.

**Methods:** We analyzed 2,923 plasma proteins in 26,690 hypertensive individuals without a prior history of AAD from the UK Biobank. LASSO and Cox regression analyses were employed to identify proteins associated with AAD, while a LightGBM algorithm was utilized to construct predictive models. Causal inference was conducted using two-sample Mendelian randomization (MR). Further mechanistic exploration included colocalization, single-cell RNA sequencing, functional enrichment, and drug-target analysis.

**Results:** Among the core hypertension proteins, we identified 186 proteins independently associated with AAD risk, with MMP12 showing the strongest association. A streamlined model, which included the top five proteins alongside age and sex, exhibited superior predictive performance (AUC: 0.791) compared to traditional risk models. MR analysis confirmed causal relationships for 21 proteins with AAD, and colocalization provided high-confidence evidence for shared genetic architecture for MMP7, CCN3, and COL6A3. Mechanistically, single-cell analysis verified cell-type-specific aortic expression of candidate genes, functional enrichment implicated extracellular matrix (ECM) pathways. Furthermore, we identified 95 FDA-approved drugs targeting 11 of these causal proteins.

**Conclusion:** This study presents the first comprehensive plasma proteomic landscape of AAD within a large hypertensive cohort, offering a high-performance predictive model, validating novel causal proteins, and identifying actionable drug targets. These findings provide crucial molecular insights into the pathogenesis of AAD and establish a solid foundation for developing early-detection strategies, improving risk stratification, and guiding precision medicine.

## Introduction

Aortic aneurysm and dissection (AAD) constitute a prevalent category of aortic pathologies. Aortic aneurysm(AA) is defined by the localized dilation of the aorta, whereas aortic dissection(AD) is characterized by the disruption of the tunica media and subsequent separation of the vessel wall^1,2^. AAD is associated with a high mortality rate, approximately 80% following rupture^3^. The pathogenesis of AAD is intricate, encompassing the degradation of elastin and collagen, phenotypic transformation of smooth muscle cells, and chronic inflammation of the aortic wall^4–7^. Additionally, fluctuations in blood pressure are believed to contribute to the development of AAD by affecting aortic dilation^8^. AAD is responsible for 2% of deaths in Western countries, and despite advancements in surgical interventions, its incidence and mortality rates remain substantial^9^.

Hypertension, a widespread cardiovascular condition, is closely linked to the occurrence of AAD. With the increasing prevalence of hypertension in recent years, the incidence of AAD has also risen, with approximately 80% of AAD patients having a history of hypertension^8,10^. This evidence indicates that the hypertension epidemic may substantially contribute to the increasing incidence of AAD. Primarily, hypertension exacerbates mechanical stress on the vascular wall, inducing structural alterations such as arteriosclerosis and decreased elasticity, thereby heightening the risk of AAD development^11^. Additionally, hypertension leads to endothelial dysfunction, which further facilitates the formation and progression of aneurysms^12^. From a genetic causality standpoint, a MR study has confirmed the causal link between hypertension and AAD, notably highlighting a significant association between elevated diastolic blood pressure and increased AAD risk^13^. Research has also identified specific genetic variants associated with susceptibility to both hypertension and cardiovascular diseases, potentially elevating AAD risk by influencing blood pressure regulation and vascular structure^12^.

However, it is particularly noteworthy that, despite this strong association, recent studies have suggested that standard antihypertensive therapy does not provide significant benefits for all AAD patients.^14,15^. This ostensibly paradoxical finding suggests that the pathogenic impact of hypertension on AAD may extend significantly beyond mere hemodynamic factors, potentially involving complex molecular biological networks or metabolic regulatory mechanisms. Despite substantial advancements in surgical and interventional treatments for AAD, early detection and pharmacological intervention in its progression among hypertensive individuals remain crucial for reducing disease burden and mortality rates. The complex interplay of genetic, environmental, and lifestyle factors has left the mechanisms linking hypertension and AAD not fully understood, necessitating an integrated approach to uncover these underlying mechanisms and develop reliable predictive tools for the disease. Proteins, as the end products of gene-environment interactions, serve as the direct effectors of biological functions.

With advancements in high-throughput proteomics, the identification of potential protein biomarkers has become a critical strategy for predicting disease risk and elucidating pathogenic mechanisms.^16–19^. Due to the minimally invasive and convenient nature of their detection, plasma proteins are the preferred choice for large-scale clinical cohort studies. Considering the well-established association between hypertension and AAD, coupled with the limitations of conventional antihypertensive therapy, the development of a specific proteomic map for AAD in the context of concurrent hypertension remains largely unexplored. Research within this high-risk population is essential, as it will facilitate the precise identification of key pathogenic proteins common to both hypertension and AAD, while also enhancing our understanding of their predictive value and underlying molecular mechanisms in disease pathogenesis. The UK Biobank Pharma Proteomics Project (UKB-PPP) has quantified nearly 3,000 proteins in over 50,000 participants using the Olink Proximity Extension Assay, thus providing a comprehensive and large-scale proteomic foundation for prospective investigations into the associations between various proteins and diseases.

It is noteworthy that, although observational studies can offer preliminary insights, they are constrained by confounding factors and the potential for reverse causality, which implies that some proteins exhibiting strong correlations may not necessarily be true pathogenic drivers^18^. In recognition of this, our study not only performed observational analyses and developed predictive models but also utilized Mendelian randomization to acquire comprehensive evidence of proteins with a genetic causal association with AAD incidence. Mendelian randomization has emerged as a method for inferring causality between an exposure and an outcome by employing genetic variants as instrumental variables. Unlike observational studies, Mendelian randomization studies are less susceptible to confounding factors and reverse causality^20^.

From a proteomics perspective, this study aims to systematically identify key plasma proteins that contribute to AAD development in a high-risk population of individuals with hypertension and to evaluate their efficacy as predictive biomarkers and potential causal mechanisms. The research methodology is as follows: Initially, we applied LASSO regression to conduct a preliminary screening of core protein features within the hypertensive cohort. Subsequently, we employed machine learning algorithms to develop an AAD risk prediction model, assessing its efficacy in risk stratification. To further substantiate causal relationships, we performed a two-sample MR analysis, incorporating cis-pQTL data from the UK Biobank-PPP and the deCODE database, alongside summary statistics from a large-scale genome-wide association study (GWAS) comprising 11,148 AAD cases and 708,468 controls. For proteins identified as candidates through causal inference, we conducted comprehensive downstream analyses, including single-cell expression localization, functional pathway enrichment, protein-protein interaction network construction, and drug target prediction. This integrative approach seeks to thoroughly elucidate the potential molecular pathological mechanisms, thereby providing a theoretical foundation for addressing the limitations of conventional antihypertensive therapy and advancing novel therapeutic strategies for AAD.

## Methods

### Population and Study Design

The UK Biobank (UKB) (https://www.ukbiobank.ac.uk/) is a population-based, prospective cohort study that recruited over 500,000 participants aged 40 to 69 between 2006 and 2010, with multiple follow-ups^21^.At baseline, participants provided sociodemographic, lifestyle, and health information, and blood samples were collected for genotyping and biochemical analysis. In this study, we included 26,690 participants who met the following criteria: they had baseline plasma protein data, were diagnosed with hypertension at baseline, had no baseline history of AAD, and had not withdrawn consent or been lost to follow-up. Proteins with a missingness rate below 20% were included. Figure 1 illustrates our study workflow, where we conducted a comprehensive proteomic study to both predict the incidence of AAD in a hypertensive population and assess the causal role of associated proteins in AAD, providing targets for understanding its underlying mechanisms.

**Fig. 1.**
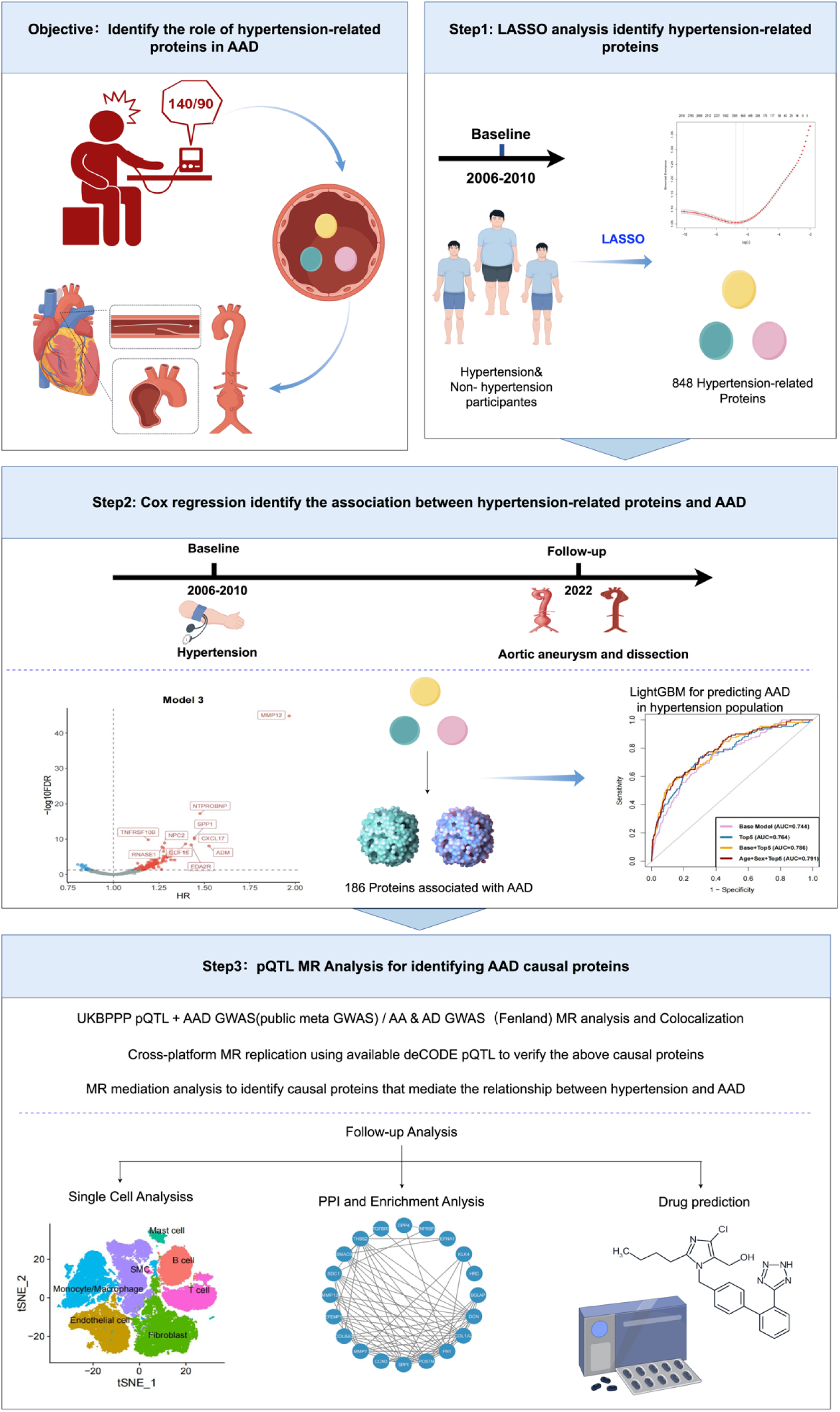
Study overview and summary. Initially, the LASSO regression technique was utilized to filter hypertension-related proteins from a pool of 2,923 plasma proteins. Subsequently, the Cox regression analysis was employed to identify proteins associated with increased risk of acute aortic dissection (AAD) within the hypertensive cohort and to develop a predictive model. Following this, Mendelian randomization was conducted to validate the causal relationships between the proteins identified through Cox regression analysis and AAD, along with its subtypes. Lastly, single-cell analysis, protein-protein interaction analysis, enrichment analysis, and drug prediction were performed on the proteins demonstrating causal relationships.

### Proteomics Analysis

Baseline blood samples were collected during the participant recruitment phase between 2006 and 2010. These samples were subsequently analyzed using the antibody-based Olink Explore Proximity Extension Assay technology for standardized quantitative detection and sample processing^22^. The complete protocols for sample pre-processing, plasma analysis using Olink proteomics technology, data processing steps, and the quality control system have been detailed in previously published literature. A total of 2,923 proteins were quantified using this platform. To ensure data quality and integrity, protein data from participants with more than 20% missing values were first excluded. For the remaining 2,911 proteins, values below the limit of detection (LOD) were imputed using the K-Nearest Neighbors (KNN) algorithm. The remaining protein levels were Z-score standardized for subsequent analysis^18^.

### Diagnosis of Hypertension and AAD

Diagnosed cases were identified through linkage to various health-related records, including hospital inpatient data, death registry records, primary care data, and self-reported information in the UK Biobank. In our study, the screening for baseline hypertension cases was based on a comprehensive assessment of several indicators: (1) a doctor-diagnosed vascular or heart problem reported as hypertension, (2) a self-reported non-cancer illness code for hypertension, (3) a diagnosis of hypertension at baseline (ICD-10: I10-I15), and (4) blood pressure measurements at baseline meeting the criteria for hypertension. This method provides a comprehensive overview of potential hypertension cases^23^. In our study, baseline AAD cases and incident cases during follow-up were identified exclusively based on the International Classification of Diseases, 10th Revision (ICD-10), primarily including codes I71.0-I71.9. The follow-up duration was calculated from the baseline date to the date of AAD diagnosis, death, or the censoring date of hospital event statistics (i.e., Hospital Episode Statistics for England: October 31, 2022; Scottish Morbidity Record for Scotland: August 31, 2022; Patient Episode Database for Wales: May 31, 2022), whichever came first.

### Covariate Assessment

In our study, we included sociodemographic, socioeconomic, and behavioral factors as covariates for analysis, including sex, age, assessment center, education level, employment, household income, Townsend Deprivation Index (TDI), smoking status, alcohol consumption, sleep duration, physical activity, and BMI. Employment status was categorized as employed or unemployed. Household income was divided into four groups: <£18,000, £18,000-£30,999, £31,000-£51,999, and >£52,000. TDI scores were divided into quartiles as a categorical variable. Smoking status was classified as current, former, or never. Alcohol consumption was categorized as never, occasional, 1-2 times per week, 3-4 times per week, or daily. Physical activity level was determined based on Metabolic Equivalent Task (MET) minutes per week for moderate activity and categorized as sufficient or insufficient. This study adopted updated cardiovascular health diet standards to classify participants’ dietary status as “adequate intake” or “inadequate intake”^24^. Sleep duration was categorized as <7, 7.0-8.0, or ≥9 hours/day. BMI was classified according to WHO standards: <18.5 kg/m², 18.5-24.9 kg/m², 25.0-29.9 kg/m², and ≥30 kg/m². Missing covariate values were handled using multiple imputation by chained equations^17^. Identification of Core Hypertension Proteins using LASSO Regression LASSO regression, by imposing an L1 regularization penalty, can shrink the coefficients of some variables to zero, thus achieving variable selection and model simplification, making it particularly suitable for high-dimensional data analysis^25^. In this study, we performed quality control on 2,923 plasma proteins from the UKB database, excluding proteins with a missing rate exceeding 20% among participants to ensure data reliability. Subsequently, we employed a LASSO regression model, using lambda.min as the optimal penalty coefficient, to screen for core proteins associated with hypertension.

### Association Analysis

We used Cox proportional hazards regression models to assess the association between protein levels and the risk of aortic dissection (AAD) and its subtypes, calculating hazard ratios (HR) and their 95% confidence intervals (CI). Three models were specified: Model 1 was a univariable analysis; Model 2 adjusted for age and sex; Model 3 further adjusted for assessment center, education, Townsend Deprivation Index, employment status, income, smoking, alcohol consumption, physical activity, healthy diet score, and BMI. Given the significant variation in AAD incidence with age and sex, we conducted subgroup analyses stratified by age (<55 vs. ≥55 years) and sex (male vs. female). All regression analyses were corrected for the false discovery rate (FDR) to control for false positives from multiple hypothesis testing, with P < 0.05 defined as statistically significant. To test the robustness of our results, we performed a series of sensitivity analyses: (1) excluding participants with a baseline history of connective tissue disease; (2) additionally adjusting for baseline history of major chronic diseases (including diabetes, hyperlipidemia, cardiovascular disease, and cancer) in the Cox model; (3) excluding participants diagnosed with AAD within two years of enrollment, as their abnormal protein levels were more likely due to undiscovered early-stage disease rather than being causal.

### Mendelian Randomization Analysis

We conducted a two-sample MR analysis using protein quantitative trait loci (pQTLs) from the UKB-PPP^22^. GWAS summary data for AAD were obtained from a publicly available meta-analysis of large-scale GWAS, including 11,148 cases and 708,468 controls of European ancestry; GWAS summary data for AA and AD were both sourced from the FinnGen database, including 9,937 cases and 463,106 controls, and 1,150 cases and 463,106 controls, respectively^26,27^. To account for potential sample overlap, this study used pQTLs from the deCODE study for replication^28^. To investigate the potential mediating role of some proteins between hypertension and AAD, we performed a mediation analysis using MR. The hypertension GWAS data were from FinnGen, including 154,630 cases and 345,634 controls^27^. When only one pQTL was available for a protein, the Wald ratio method was applied. For proteins with two or more genetic instruments, a fixed- or random-effects inverse-variance weighted (IVW) model was used, considering potential heterogeneity. Before conducting the MR analysis, we followed these steps: First, pQTLs were pruned for linkage disequilibrium (LD) at a genome-wide significance threshold of P < 5E-8 (based on the 1000 Genomes European reference panel), ensuring an LD r² < 0.001 and a clumping distance of 10,000 kb. Then, instruments with an F-statistic > 10 were selected to test the statistical power of each pQTL, classifying them as adequate. To avoid reverse causality, we applied Steiger filtering to remove variants more strongly associated with the outcome than the exposure^29^. R version 4.3.2 with the R package TwoSampleMR (version 0.5.7) was used for all statistical analyses. Cochran’s Q test was performed to assess heterogeneity^30^. The MR-Egger regression intercept was used to evaluate potential horizontal pleiotropy^31^. Results are presented as odds ratios (OR) and 95% confidence intervals (CI).

### Colocalization Analysis

For proteins showing significant association with AAD in both the cohort and MR analyses, we performed colocalization analysis using the coloc R package with default prior probabilities of p1=1E-4, p2=1E-4, and p12=1E-5^29^. Colocalization assesses the probability that two traits share the same causal variant, rather than sharing a variant by coincidence due to LD. This analysis is crucial for strengthening the evidence of a causal relationship between candidate proteins identified through MR analysis and AAD. A posterior probability (PP.H4) greater than 0.60 was used as the threshold for determining significant colocalization.

### Cell-Type Expression Analysis

To evaluate the target genes corresponding to plasma proteins causally related to AAD, we analyzed their cell type-specific expression based on a single-cell RNA sequencing dataset of aortic tissue from AAD cases and controls (GSE226492) ^32^. Using the “Seurat” package, we performed data preprocessing and transformation^33^. We removed genes with fewer than 3 counts in a cell and cells with fewer than 300 unique feature counts. At the cell level, we retained cells with detected gene counts between 500 and 5000, mitochondrial gene percentage <10%, ribosomal gene percentage >3%, and red blood cell gene percentage <0.1%. For normalization, we used the NormalizeData function, followed by FindVariableFeatures to identify highly variable genes and ScaleData to scale all genes. We then performed Principal Component Analysis (PCA) and t-Distributed Stochastic Neighbor Embedding (t-SNE) for dimensionality reduction. To identify differentially expressed genes, we used the FindAllMarkers function with the control group as a baseline. The criteria for differential expression were an average log-fold change (|log₂FC|) > 0.25 and an FDR-adjusted p-value < 0.05.

### Predictive Model Construction

The dataset was randomly split into a training set (70%) and a testing set (30%) using stratified sampling. We evaluated five machine learning models (LightGBM, XGBoost, Random Forest, SVM, GLM) for predicting AAD incidence. The Light Gradient Boosting Machine (LGBM) model, which showed the best performance, was selected for subsequent analysis. We ranked proteins based on their contribution to the model’s predictive performance using SHAP (SHapley Additive exPlanations) values and visualized the selected proteins with SHAP plots^34,35^. We then compared the predictive performance of models built with the top 10, top 5, and top 3 proteins. We also compared our top-5 protein model with a previously published AAD protein prediction model^36^. Additionally, we compared the performance of our protein-based model with a model based on traditional risk factors (age, sex, ethnicity, education, TDI, BMI, physical activity, smoking, and alcohol). Finally, we combined the protein and traditional models to observe the improvement in predictive performance. These analyses were conducted using the shapviz, lightgbm, xgboost, and caret packages.

### Enrichment and Protein-Protein Interaction Analysis

To systematically elucidate the functions and interactions of the selected proteins, we constructed a protein-protein interaction (PPI) network using the STRING database (https://string-db.org/)^37^. Subsequently, we used the clusterProfiler package (version 4.4.4) for Gene Ontology (GO) functional annotation and Kyoto Encyclopedia of Genes and Genomes (KEGG) pathway enrichment analysis^38^.

### Drug Prediction

To explore the clinical translational potential, the Drug-Gene Interaction Database (DGIdb, https://dgidb.org/) was used to screen for potential drugs targeting the core genes, aiming to provide new strategies for the prevention or treatment of AAD in hypertensive patients^39^.

## Results

### 1. Population Characteristics

Following the exclusion of 73 individuals with baseline AAD, 3,579 individuals of non-European ancestry, and 114 individuals lost to follow-up, our study comprised 49,247 participants of European ancestry. At baseline, the cohort included 26,690 individuals diagnosed with hypertension (mean age: 59.37 years) and 22,557 individuals without hypertension (mean age: 54.42 years). Over a median follow-up period of 13.6 years (interquartile range: 12.9-14.3), amounting to 647,849 person-years, we documented 493 incident cases of AAD, representing 1.00% of the total sample. These included 35 cases of AD and 475 cases of AA. Specifically, within the hypertensive cohort, 370 AAD events were recorded (AD: 25; AA: 356), whereas the non-hypertensive cohort experienced 123 AAD events (AD: 10; AA: 119). The incidence of AAD was significantly elevated in the hypertensive cohort, as evidenced by Pearson’s Chi-squared test (χ² = 89.757, p-value < 2.2e-16; refer to Supplementary Table 1). Supplementary Table 2 provides the baseline characteristics of the hypertensive cohort. Overall, individuals who developed AAD exhibited significant differences from those who did not, particularly in terms of age, sex, smoking status, income, and education. Those who developed AAD tended to be older, male, have a history of smoking, possess vocational education, and have lower household incomes.

### 2. Screening of Core Proteins for Hypertension

Out of the initial 2,923 plasma proteins, those with more than 20% missing values were excluded, resulting in a dataset of 2,911 proteins for further analysis involving 26,690 hypertensive patients and 22,557 control subjects. Employing a LASSO regression model, we identified 848 core proteins that exhibited significant associations with hypertension (refer to Supplementary Fig. 1A, B). KEGG pathway enrichment analysis of these core proteins indicated significant enrichment in pathways such as cytokine-cytokine receptor interaction, PI3K-Akt signaling pathway, cell adhesion molecules, and ECM-receptor interaction (see Supplementary Fig. 1C). Subsequent Gene Ontology (GO) analysis demonstrated that at the cellular component (CC) level, these proteins were predominantly localized within the collagen-containing extracellular matrix. At the biological process (BP) level, they were primarily involved in processes such as chemotaxis, cell chemotaxis, and cell-matrix adhesion. At the molecular function (MF) level, these proteins were chiefly associated with glycosaminoglycan binding (refer to Supplementary Fig. 1D).

### 3. Identification of AAD-Associated Proteins among Core Hypertension Proteins

Among the 848 core proteins associated with hypertension, univariable Cox regression analysis (Model 1) identified 391 proteins as significantly correlated with the risk of acute aortic dissection (AAD) (refer to Fig. 2A and Supplementary Table 4). Upon adjusting for age and sex in Model 2, 266 proteins maintained their significant association (see Fig. 2B and Supplementary Table 5). Further adjustment for multiple covariates in Model 3 revealed that 186 proteins continued to exhibit an independent association with AAD (refer to Fig. 2C and Supplementary Table 6). Notably, across all three models, Matrix Metalloproteinase-12 (MMP12) exhibited the most pronounced association with AAD. Additionally, proteins such as CXCL17, GDF15, NPC2, NT-proBNP, and TNFRSF10B consistently demonstrated significant associations across the models. Sensitivity analyses, based on Model 3, were performed to enhance robustness. By excluding cases that occurred within two to ten years post-enrollment, 161 and 10 proteins, respectively, remained significantly associated, indicating their potential relevance to the long-term progression of the disease (Fig. 2F, G, Supplementary Tables 9, 10). Subsequent analysis using unimputed data corroborated a significant association for 188 proteins (Fig. 2H, Supplementary Table 11).

**Fig. 2.**
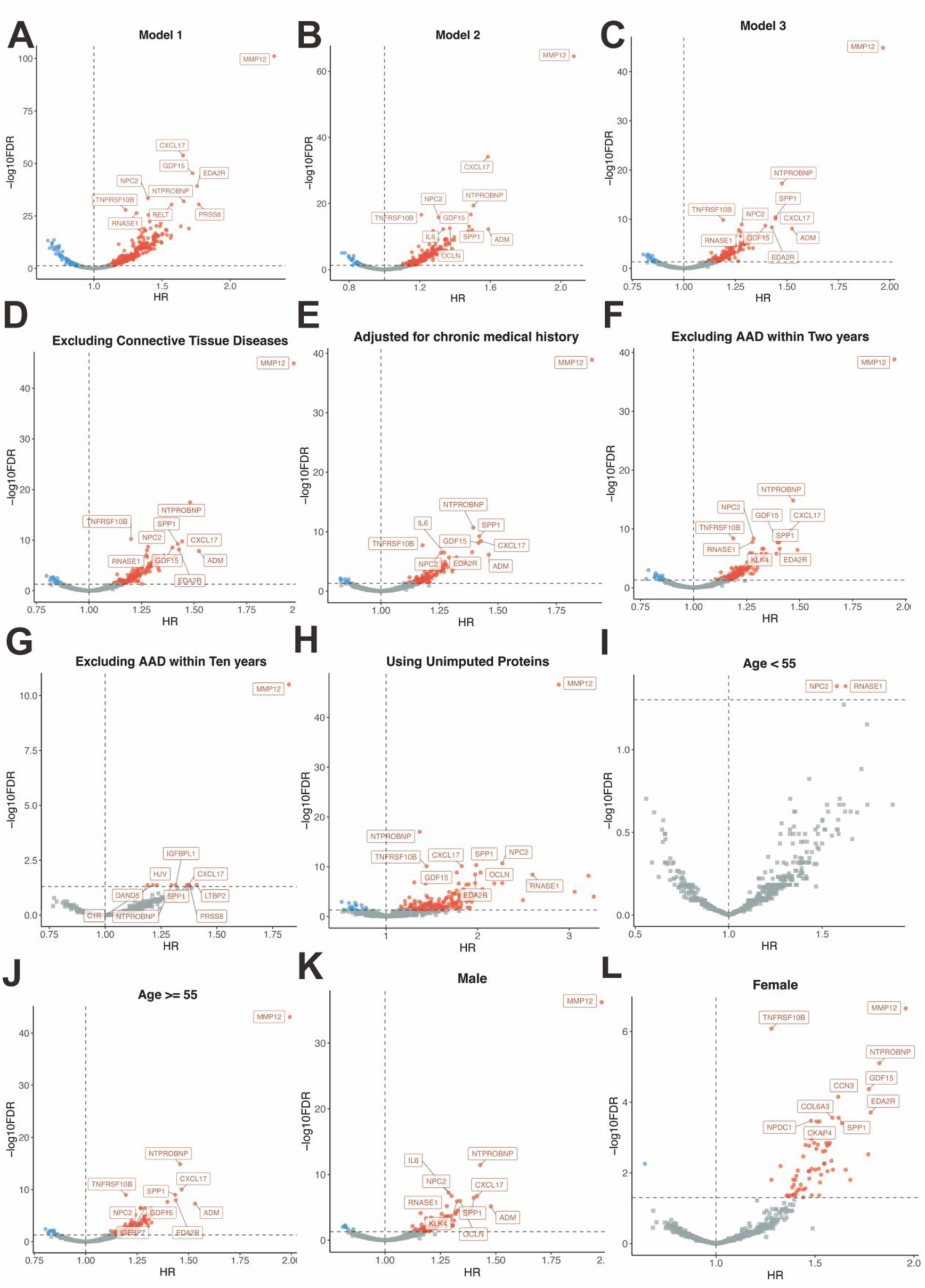
Volcano Plot of the Prospective Association of Proteins with Aortic Aneurysm and Dissection. (A-C) Construction of three sequential multivariable Cox proportional hazards models to estimate the hazard ratios (HRs) of proteins for AAD risk. The models were progressively adjusted as follows: Model 1, unadjusted; Model 2, adjusted for age and sex; Model 3, further adjusted for assessment center, educational attainment, Townsend deprivation index, employment status, income, smoking, alcohol consumption, physical activity, healthy diet score, and BMI. (D-H) Sensitivity analyses based on Model 3: (D) excluding individuals with connective tissue diseases at baseline; (E) additionally adjusting for history of chronic diseases; (F) excluding cases occurring within two years of baseline; (G) excluding cases occurring within ten years of baseline; (H) using unimputed protein data. (I-L) Subgroup analyses stratified by (I, J) age and (K, L) sex, using Model 3 as the base model. All P values were corrected for multiple testing using FDR method. FDR false discovery rate, HR hazard ratio.

Subgroup analyses revealed that the number and strength of associations between plasma proteins and AAD were more pronounced in individuals aged 55 years and older, as well as in male participants, aligning with the epidemiological patterns of AAD (Fig. 2 I-L, Supplementary Tables 12-15). In the sex-stratified analysis, 39 proteins were significantly associated in both male and female groups. Notably, while MMP12 emerged as the top protein in most subgroups, NPC2 and RNASE1 were most significantly associated with AAD in individuals under 55 years of age.

### 4. Construction of a Machine Learning-Based Prediction Model

In the hypertensive cohort, five machine learning models were developed, with the LightGBM (LGB) model exhibiting superior predictive performance, achieving an Area Under the Curve (AUC) of 0.780 (95% CI: 0.733–0.829) in the validation set (Supplementary Table 16, Fig. 3A). SHAP analysis identified the top 10 most important proteins for prediction: MMP12, FGFBP1, CXCL17, TNFRSF10B, MB, ELN, NT-proBNP, LEP, SUSD5, and PRSS8 (Fig. 3B, C, Supplementary Table 17). Models built with the top 10, top 5, and top 3 proteins yielded AUCs of 0.771, 0.764, and 0.744, respectively (Supplementary Table 18, Fig. 3D). Our model outperformed a previously published protein panel (AUC 0.733, 95% CI:0.684-0.783) (Supplementary Table 18, Fig. 3E). The Top-5 protein model was selected as the optimal model.

**Fig. 3.**
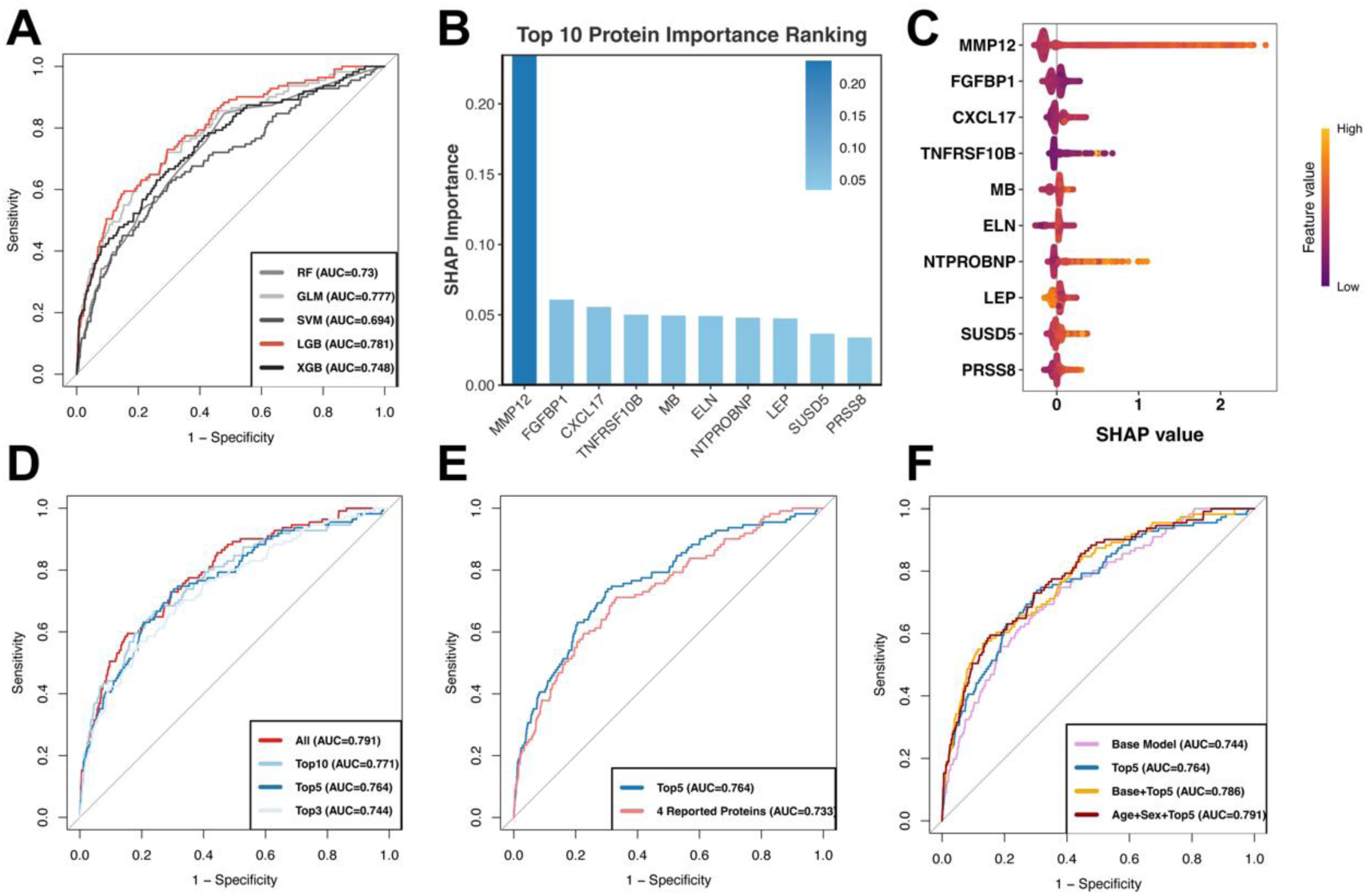
Machine learning models for AAD risk prediction and key protein biomarkers in hypertension. (A) Comparison of predictive performance (AUC) among different machine learning models in the validation set. (B) Ranking of the top 10 key protein biomarkers based on SHAP values. (C) SHAP swarm plot showing the importance of protein biomarkers for AAD prediction. (D) Predictive performance of models constructed with Top3, Top5, and Top10 protein biomarkers. (E) Comparison of predictive performance between the optimal model in this study and the protein panel from a previous study. (F) Predictive performance of the Top5 protein biomarkers combined with traditional factors (full traditional factors vs. age + gender). SHAP SHapley Additive exPlanations, AUC Area Under the Curve, RF Random Forest, GLM Generalized Linear Model, SVM Support Vector Machine, LGB Light Gradient Boosting Machine, XGB eXtreme Gradient Boosting.

Combining the Top-5 proteins with traditional risk factors improved the AUC to 0.786 (95% CI: 0.741-0.830), which was superior to the model with traditional factors alone (AUC: 0.744, 95% CI: 0.699-0.790). Interestingly, a simplified model combining the Top-5 proteins with only age and sex achieved an even higher AUC of 0.791 (95% CI: 0.748-0.834), suggesting that a more parsimonious model could enhance predictive accuracy (Supplementary Table 18, Fig. 3F).

### 5. Mendelian Randomization and Colocalization Analysis

Of the 186 proteins associated with AAD, cis-pQTL data were available for 167 from the UKB-PPP. A two-sample MR analysis identified eight proteins with a significant causal association with AAD (P < 0.05). Colocalization analysis provided robust evidence that MMP7 and CCN3 share a causal variant with AAD, as detailed in Supplementary Table 19 and illustrated in Figure 4A. Furthermore, 13 proteins were identified as causally associated with AA, with CCN3 exhibiting colocalization, while 7 proteins were linked to AD, with COL6A3 showing colocalization, as presented in Supplementary Table 19 and Figure 4A. The MR-Egger intercept test did not detect any significant horizontal pleiotropy (Supplementary Table 19).

**Fig. 4.**
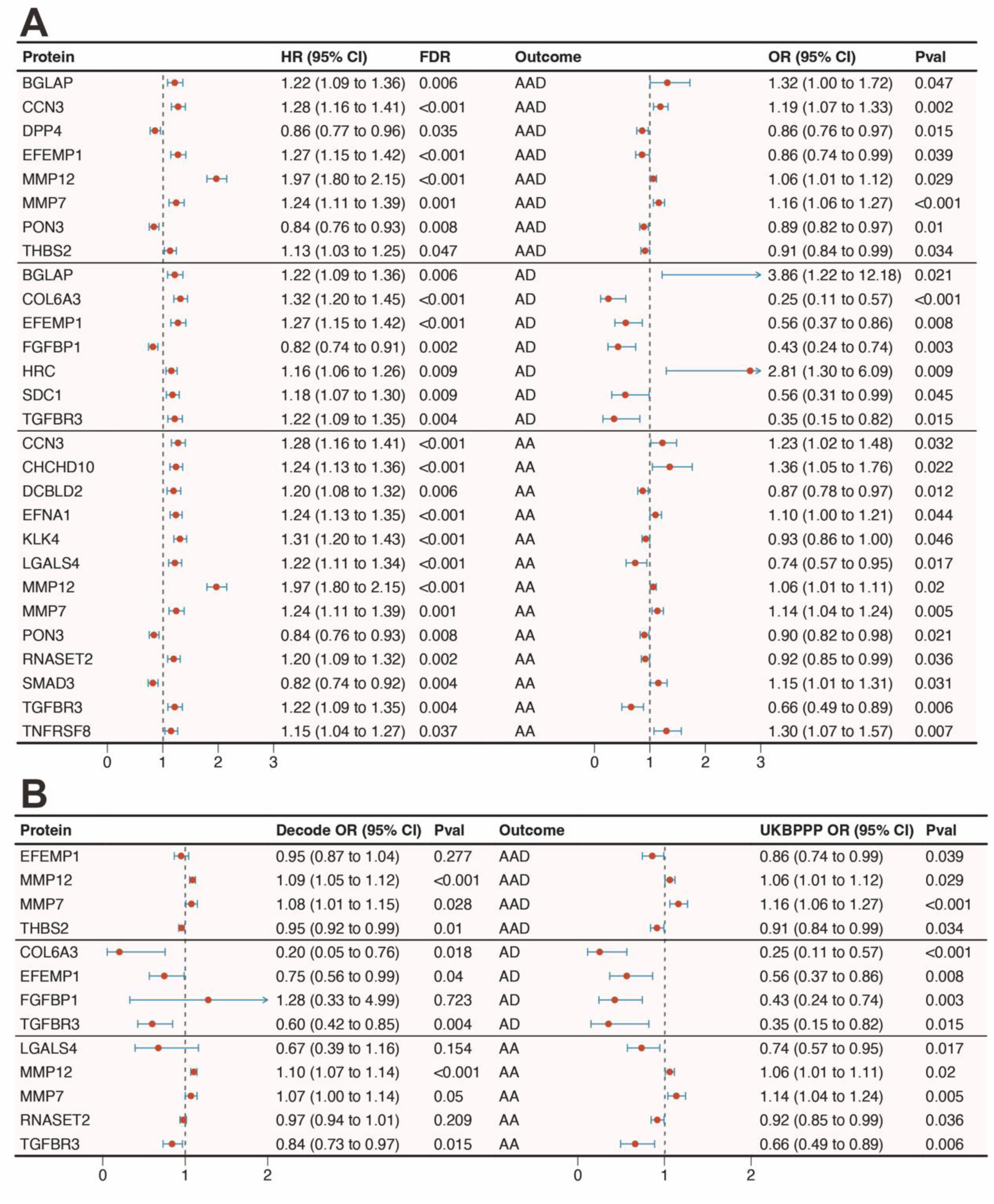
Mendelian randomization analyses for causal associations between plasma proteins and AAD and its subtypes. (A) The forest plot presents the HRs from the observational analysis alongside the causal estimates from MR. The MR analysis was conducted employing pQTL data sourced from the UKB-PPP. (B) The forest plot presents the results of the replication MR analysis, which utilized pQTL data from deCODE. All MR estimates were derived from the IVW method. For each protein, the forest plot displays the OR and 95% CI for the risk of AAD and its subtypes associated with a genetically predicted 1-SD increase in plasma protein level. HR Hazard Ratio, OR, Odds Ratio, CI Confidence Interval, IVW Inverse-variance Weighted, UBPPP UK Biobank Pharma Proteomics Project.

Replication MR analysis using cis-pQTLs from the deCODE database corroborated the direction of effect for most proteins and retained significance for several key proteins, including MMP12, MMP7, and THBS2 for AAD, as well as COL6A3, EFEMP1, and TGFBR3 for AD, and MMP12 and TGFBR3 for AA (Supplementary Table 20, Figure 4B). MR analysis further established a causal relationship between hypertension and AAD (Supplementary Table 21).

Mediation analysis indicated that MMP7 and PON3 significantly mediate the effect of hypertension on AAD, with higher MMP7 expression increasing AAD risk (mediation proportion: 3.35%) and lower PON3 expression offering a protective effect (mediation proportion: 2.09%) (Supplementary Table 21).

### 6. Single-Cell Type Expression Analysis of Candidate Protein-Encoding Genes

We conducted an analysis of the expression of protein-coding genes with established causal links to AAD within a human aortic single-cell RNA-seq dataset. Aortic cells were categorized into seven primary types (Fig. 5A). Among the 21 genes examined, CCN3, predominantly expressed in fibroblasts and smooth muscle cells, and SMAD3, primarily expressed in endothelial cells, were found to be significantly upregulated in AAD patients compared to controls (Fig. 5B-C). In contrast, EFEMP1, THBS2, COL6A3, SDC1, TGFBR3, and RNASET2 were significantly downregulated in their respective enriched cell types, aligning with the findings from the MR analysis (Supplementary Table 22, Fig. 5B-C).

**Fig. 5.**
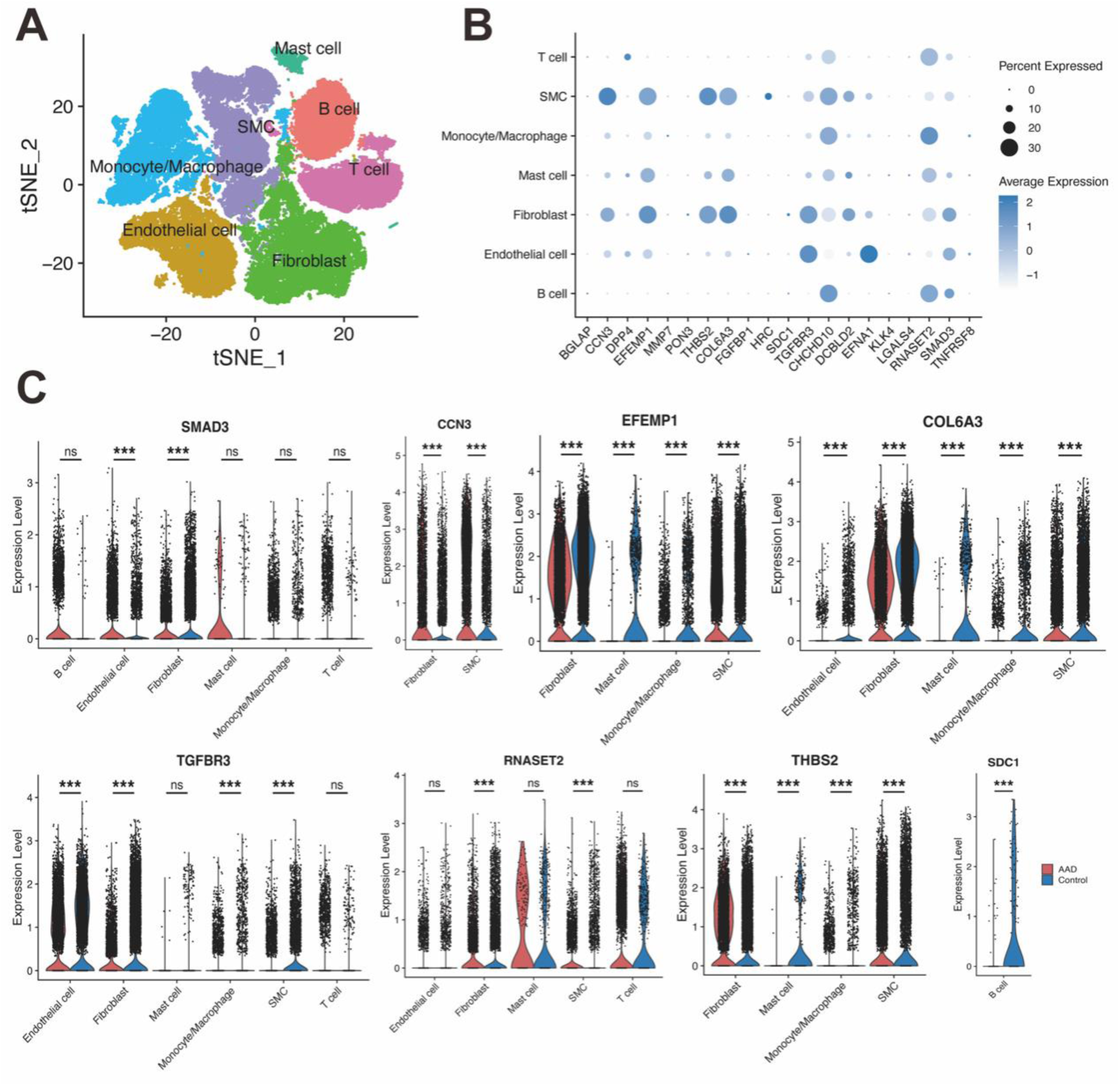
Single-Cell Transcriptomic Profiling of Causal Protein-Encoding Genes in the Human Aorta. (A) t-SNE visualization depicting the seven major cell types identified in human aortic tissue from a published single-cell RNA-seq dataset. (B) Bubble plot displaying the cell type-specific enrichment and average expression levels of the protein-encoding genes across the defined aortic cell populations. (C) Violin plots comparing the expression levels of selected differentially expressed genes between aortic samples from AAD patients and controls within their primary enriched cell types. Differential expression analysis was performed using the Wilcoxon rank-sum test. All tests were two-sided and adjusted for multiple comparisons. Significance was defined as |log₂FC| > 0.25 and FDR < 0.05. Asterisks denote statistical significance levels: *FDR < 0.05, **FDR < 0.01, ***FDR < 0.001,ns FDR>=0.05.

### 7. Protein-Protein Interaction, Enrichment, and Drug Prediction Analysis

A PPI network constructed from the 21 causal proteins demonstrated a highly significant, non-random interaction network (PPI enrichment p-value < 1.0e-16), indicating potential functional synergy (Supplementary Table 23, Supplementary Fig. 2A). KEGG pathway analysis revealed enrichment in the “ECM-receptor interaction” and “Focal adhesion” pathways, primarily driven by COL6A3, THBS2, and SDC1 (Supplementary Table 24, Supplementary Fig. 2B). GO analysis underscored processes associated with extracellular matrix organization, including components such as collagen-containing ECM, and molecular functions like heparin binding and serine-type endopeptidase activity (refer to Supplementary Table 25 and Supplementary Figure 2C).

Drug-gene interaction analysis conducted via the DGIdb identified 11 out of 21 proteins as ‘druggable’ targets, revealing 376 potential interactions with 95 FDA-approved drugs. Notably, the antibody-drug conjugate Indatuximab ravtansine, which targets SDC1, demonstrated the highest interaction score. This analysis also uncovered intricate interactions between commonly used antihypertensive medications and proteins associated with AAD. Specifically, the angiotensin-converting enzyme (ACE) inhibitor Captopril was predicted to interact with both the risk-associated MMP12 and the protective DPP4. Additionally, several drugs from alternative pathways were identified for their potential roles in inflammation suppression and calcium metabolism. These include Zileuton and Doxycycline, which target MMP family proteins, as well as various calcium metabolism modulators (e.g., Paricalcitol, Calcitonin) that regulate BGLAP.

## Disscusion

Hypertension is a major contributor to various cardiovascular diseases; however, its molecular mechanisms in acute aortic dissection (AAD) remain largely elusive and challenging to predict at the population level^40^. By integrating large-scale proteomics data from a prospective cohort with genetic analysis, this study systematically investigates plasma protein biomarkers associated with the development of AAD in a hypertensive population. This research offers an innovative and comprehensive perspective for understanding, predicting, and addressing AAD development within this high-risk group. We have developed a multi-dimensional research framework that encompasses biomarker screening, mechanistic exploration, and clinical prediction.

Initially, we identified 848 core proteins associated with hypertension and elucidated their enriched biological functions. Both KEGG and GO enrichment analyses revealed that these proteins are linked to pathways involving the ECM, including ECM-receptor interaction (KEGG), collagen-containing ECM (GO Cellular Component), and cell-matrix adhesion (GO Biological Process). During the progression of hypertension, Ang II facilitates ECM turnover, resulting in increased vascular ECM stiffness and diminished compliance to tensile forces from blood flow, which subsequently exacerbates the hypertensive condition^41,42^. Among the proteins analyzed, 186 were significantly associated with the risk of AAD after adjusting for variables such as sex, age, assessment center, education, employment, household income, Townsend Deprivation Index, smoking status, alcohol consumption, sleep duration, physical activity, and BMI.

In terms of clinical prediction, MMP12, FGFBP1, CXCL17, TNFRSF10B, and myoglobin (MB) were identified as key factors in our predictive model, highlighting their potential for clinical application. Research has demonstrated that MMP12, a matrix metalloproteinase, can degrade ECM components, thereby impacting the structural integrity and function of the vascular wall^43^. In the pathological context of AAD, interleukin-3 (IL-3) can significantly enhance the expression and activity of macrophage-derived MMP12, which is closely associated with degenerative changes and the inflammatory response of the vascular wall^43,44^. Moreover, numerous studies have documented that plasma concentrations of MMP12 are markedly elevated in patients with Stanford Type A AD, aligning with the findings of our research^36,45,46^. FGFBP1 is intricately linked to blood pressure regulation and plays a role in the pathogenesis of hypertension^47,48^. CXCL17, identified as a mucosal chemokine, has been associated with various cancers according to current research^49,50^. Additionally, as a chemokine, CXCL17 is involved in the inflammatory processes of multiple organs, potentially indicating a connection to the vascular inflammation observed in AAD^51,52^. TNFRSF10B, a member of the death receptor family, is capable of mediating myocardial cell apoptosis via the TRAIL-DR5 signaling pathway^53^. In parallel, our study identified elevated blood myoglobin (MB) levels as a significant risk factor for AAD. While MB is commonly regarded as a diagnostic marker for acute coronary syndrome, indicative of potential cardiomyocyte lysis, its elevation in other diseases does not invariably originate from cardiomyocytes^54^. The source of MB in AAD, as observed in our study, warrants further investigation. We evaluated the predictive capabilities of the top proteins identified from our hypertension-focused screening against a four-protein AAD prediction panel from a prior study, which was not specifically selected for a hypertension cohort. Our findings indicated that the Top-3 protein signature exhibited superior predictive performance within the hypertensive cohort compared to the aforementioned four-protein panel^36^. This suggests that concentrating on hypertension-related proteins enhances our understanding of the comorbidity relationship and augments predictive value. Notably, 80% of AAD patients present with underlying hypertension, highlighting the importance of directing primary screening efforts towards this high-risk group to optimize the allocation of medical resources and achieve maximal predictive efficacy^23^.

Additionally, our observational analyses indicated that the associations between plasma proteins and AAD differed across various subgroups. Although MMP12 demonstrated the strongest association in most analyses, NPC2 and RNASE1 emerged as the most significant proteins in the younger (<55 years) subgroup, indicating their potential utility in predicting AAD among younger hypertensive patients. In female participants, the association with TNFRSF10B was notably stronger, suggesting potential sex-specific protein differences. While AAD is generally more prevalent in older male populations, its occurrence in younger and female individuals is often attributed to genetic factors or congenital cardiovascular diseases^55–58^. Our findings, which delineate distinct protein profiles across age and sex, may provide a foundation for stratified screening strategies. In light of this, we developed a model that integrates our top five proteins with age and sex, thereby simplifying the model by omitting complex covariates while achieving optimal predictive performance. This underscores the importance of customizing protein-based predictions according to key demographic factors. However, the smaller sample sizes for younger and female participants necessitate further validation of these findings.

Although the proteins identified by the machine learning predictive model are significant for predicting AAD in the hypertensive population, results from observational analyses are often vulnerable to various confounding factors and reverse causality, and thus may not adequately determine whether a protein is a true causal factor. In this study, a two-sample MR analysis was conducted to further validate the 186 identified proteins, ultimately identifying 21 proteins with a potential causal relationship with AAD and its subtypes. Specifically, the MR results indicated that 8, 7, and 12 proteins exhibited causal associations with AAD, AD, and AA, respectively. This suggests that variations in the expression levels of these plasma proteins differ across the overall disease and its various subtypes. For these proteins, colocalization analysis was further employed to ascertain whether their protein levels and disease risk are influenced by the same causal variant. High-confidence colocalization evidence was found for MMP7 and CCN3 with AAD. Additionally, CCN3 demonstrated high colocalization evidence with AA, while COL6A3 showed high colocalization evidence with AD. These findings indicate that alterations in the levels of different proteins have varying impacts on susceptibility to different manifestations of AAD. Furthermore, mouse studies have demonstrated that MMP7 transcriptionally upregulates MMP2, which subsequently mediates AngII-induced hypertension^59^. Moreover, MMP2 not only contributes to increased blood pressure but is also significantly concentrated in the aortic wall, where it plays a role in the degradation of the wall matrix, thereby having a well-documented influence on AAD formation^59^. This evidence indicates that MMP7 may serve as a potential mediator of AAD under hypertensive conditions, corroborating the findings of our mediation analysis. The CCN protein family is implicated in various cellular processes, including apoptosis, senescence, ECM production, differentiation, and proliferation. Notably, CCN3, a brain-derived bone factor, has been recently identified as a promoter of bone growth^60,61^. However, in the context of aortic health, previous research has suggested that CCN3 acts as a protective factor against AAD^62^. Contrary to this, our study’s observational and MR analyses do not support this protective role; instead, they suggest that CCN3 is more likely to be a contributing factor in the development of AAD. This discrepancy necessitates further investigation. COL6A3 is a member of the collagen family of proteins and constitutes the α-3 chain of type VI collagen, a crucial component of the extracellular matrix that plays a role in structural support and signal transduction^63^. The C-terminal cleavage product of COL6A3, known as endotrophin, is hypothesized to contribute to coronary heart disease upon its release into the bloodstream, indicating that the degradation of COL6A3 itself may be a fundamental cause of various cardiovascular diseases. This hypothesis is consistent with our MR analysis and single-cell sequencing results, which identified COL6A3 as a protective factor against AD However, our observational study reported a HR greater than 1 for COL6A3. This finding could be attributed either to elevated levels of its degradation products in the bloodstream or to a compensatory upregulation of the COL6A3 protein itself^64^. In our mediation analysis, in addition to identifying MMP7 as a mediator between hypertension and AAD, PON3 was also identified as a negative mediator between these conditions. PON3 is a member of the Paraoxonase gene family, whose members have been demonstrated to exert antioxidant effects in the pathophysiology of coronary heart disease^65^. The protective function of PON3 in AAD may be attributed to its antioxidant properties, as PON3 is capable of mitigating mitochondrial superoxide production, thereby safeguarding cells against oxidative stress-induced damage. Various mechanisms underlying AAD pathogenesis, such as hypertension and elevated metalloproteinase activity, are intrinsically associated with oxidative processes^66,67^.

The results from enrichment analysis of these proteins suggest a significant involvement of the ECM and cytoskeleton in AAD development. This aligns with the enrichment findings for hypertension-associated proteins, collectively indicating that an intact ECM and vascular wall cytoskeleton constitute the structural foundation of a healthy aortic wall. Alterations in their protein expression may represent a potential mechanism by which hypertension induces ECM modifications in AAD. Concurrently, we identified three proteins—COL6A3, THBS2, and SDC1—enriched within the ECM-receptor interaction pathway, which also demonstrate strong interactions within the PPI network. Notably, THBS2 is an adhesive glycoprotein that facilitates cell-cell and cell-matrix interactions. SDC1 is a cell-surface proteoglycan that carries both heparan sulfate and chondroitin sulfate, playing a crucial role in linking the cytoskeleton to the interstitial matrix. This suggests that the extracellular matrix and cell junctions warrant further investigation^68,69^. To explore the potential expression patterns of proteins implicated in AAD within aortic tissue, this study analyzed a publicly available single-cell sequencing dataset of whole aortic tissue. Notably, the expression levels of the two MMP family proteins examined in this study were found to be low in aortic tissue, with MMP12 being undetectable due to its extremely low expression level. Consistent with previous studies, MMP12 mRNA did not exceed the detection threshold in the aorta. However, at the protein level, an increase in MMP12 has been confirmed in the aortic wall of AAD patients. This indicates that the pro-pathogenic roles of MMP12 and MMP7 in AAD are more likely to be of hematogenous origin^70,71^. Eight proteins—SMAD3, CCN3, EFEMP1, COL6A3, TGFBR3, RNASET2, THBS2, and SDC1—exhibited differential expression between the two phenotypes (AAD versus control). The direction of this differential expression aligned with the OR derived from the MR analysis. This finding not only reinforces the evidence supporting these proteins as pathogenic factors in AAD but also provides valuable insights for future mechanistic studies and drug target development.

Our analysis offers a crucial molecular explanation for the ongoing clinical debate regarding the role of ACE inhibitors (ACEIs) in AAD progression, specifically whether they are protective or detrimental^72,73^. Previous observational studies have yielded conflicting results, with some suggesting that mere blood pressure reduction does not mitigate AAD risk^14^. We are the first to present a robust mechanistic explanation at the level of protein interactions: ACEIs manifest a dual effect. On one hand, their inhibition of the risk protein MMP12 substantiates a beneficial effect. Conversely, their off-target inhibition of the key protective protein DPP4 may underlie the potential adverse effects. We propose that the clinical net effect of any ACEIs results from a balance between its beneficial and adverse effects. This shifts the clinical inquiry from whether ACEIs should be employed to determining which antihypertensive is most appropriate for specific patient populations. Our finding that DPP4 serves as a protective protein against AAD carries significant clinical implications, particularly regarding the widely prescribed gliptin class of anti-diabetic medications^74^. The therapeutic action of these drugs, which involves systemic DPP4 inhibition, may inadvertently undermine the aorta’s intrinsic protective mechanisms, potentially increasing the long-term risk of AAD in patients with concurrent hypertension and diabetes. This observation is especially significant given the well-documented “reverse protective” effect, wherein diabetes itself acts as a protective factor against AAD^75–77^. Although the mechanism underlying this protection has remained unclear, our research introduces a novel mechanistic hypothesis: DPP4 may play a crucial role in mediating this “diabetic protective effect.” Therefore, the use of DPP4 inhibitors for managing hyperglycemia could paradoxically negate the inherent aortic protection conferred by diabetes. This highlights the critical need to elucidate the specific role of DPP4 in maintaining aortic wall homeostasis.

Beyond this, our drug screening has identified several highly promising therapeutic opportunities. Notably, the asthma medication Zileuton, through its inhibition of MMP12, may possess cross-disease therapeutic potential. Additionally, we have confirmed at the plasma protein level that doxycycline’s efficacy in limiting AAD progression is mediated by its targeting of MMP7, a key plasma target identified in our study, which is consistent with previous animal and clinical research^78,79^. Finally, the identification of BGLAP (osteocalcin) and its interactions with calcium-regulating drugs (e.g., paricalcitol, calcitonin) strongly suggests a pathological interplay among dysregulated calcium homeostasis, vascular calcification, and the compromise of aortic wall integrity, offering valuable insights for exploring AAD prevention from a metabolic standpoint.

As the first study to provide a systematic proteomic perspective on AAD pathogenesis within a hypertensive population, this research establishes a comprehensive framework that spans from causal genetics to potential clinical applications for AAD drug intervention. However, several limitations must be considered. Firstly, the UK Biobank cohort predominantly consists of middle-aged and older individuals of European ancestry, necessitating further validation to ascertain the generalizability of our findings across diverse ethnic groups and age ranges. Secondly, certain proteins were excluded from the Mendelian Randomization (MR) analysis due to the absence of robust instrumental variables, and the reliance on single-timepoint proteomic data limits the ability to capture temporal dynamic changes. Additionally, technical constraints associated with the assay may introduce a degree of uncertainty. Thirdly, our machine learning model requires validation through independent prospective clinical cohorts to evaluate its practical clinical utility and cost-effectiveness. Lastly, and most importantly, our mechanistic and drug-related conclusions are derived from computational predictions. The specific molecular mechanisms, as well as the actual efficacy and safety of the proposed drugs, necessitate confirmation through subsequent laboratory experiments and clinical trials.

In conclusion, this study employed large-scale proteomics to systematically identify a cohort of plasma proteins within a hypertensive population that exhibit a strong association with the risk of acute aortic dissection (AAD). We established and validated a causal relationship for several of these proteins with AAD and its subtypes. Additionally, we developed and validated a high-precision, protein-based predictive model for AAD, elucidating potential mechanisms through which hypertension influences AAD pathogenesis via specific proteins. These findings not only enhance our understanding of AAD pathogenesis but also provide essential biomarkers and a theoretical framework for the development of future strategies aimed at early warning, risk stratification, and targeted therapy, while also suggesting potential drug candidates for managing these comorbidities. Ultimately, our results offer novel insights into the clinical prediction and targeted treatment of AAD, underscoring the potential of proteomics-driven precision medicine to advance the management of AAD in hypertensive individuals.

## Data availability

Proteomic data were obtained from the UK Biobank resource, which supports health-related research in the public interest (https://www.ukbiobank.ac.uk/enable-your-research/register). This study utilized the UK Biobank under application number 95608. pQTL data were retrieved from the UK Biobank Pharmacogenomics Project (UKB-PPP) (https://www.synapse.org/Synapse:syn51364943) and the Icelandic Database (https://www.decode.com/summarydata/). GWAS summary statistics for AAD are available at the National Genomics Data Center (NGDC), China National Center for Bioinformation (CNCB) (OMIX accession: OMIX00 7191; https://ngdc.cncb.ac.cn/omix). GWAS summary statistics for AD, AA, and hypertension can be accessed through the FinnGen R12 study (https://www.finngen.fi/fi).

## Availability of data and materials

The datasets analyzed in this study are from: 1. UK Biobank (https://www.ukbiobank.ac.uk/); 2. Icelandic database (https://www.decode.com/sum marydata/) (accession nos. PMID: 34857953); 3. FinnGen consortium (https://www.finngen.fi/en/access_results) (accession nos. finngen_R11_T2D; finngen_R11_I9_CHD).

## Acknowledgements

We would like to express our sincere gratitude to all staff members of the UK Biobank for their valuable contributions to this study, as well as to the researchers and all participants of the Icelandic Database and FinnGen study for making their summary data publicly available. We also appreciate the language polishing service offered by Home for Researchers (www.home-for-researchers.com) that enhanced the clarity and fluency of the manuscript, and thank Figdraw (www.figdraw.com) for its technical assistance in creating graphical abstracts and flowcharts.

## Author contribution

Jiaqi Hou: Conceptualization, Methodology, Formal analysis, Software, Writing—original draft. Longjiang Wu: Methodology, Investigation, Formal analysis, Visualization, Writing—original draft. Lihua Lin: Validation, Data curation, Formal analysis, Writing—review & editing. Meichen Pan: Validation, Investigation, Data curation. Jing Huang: Validation, Writing—review & editing. Jianghua Du: Validation, Data curation. Shujuan Wang: Investigation, Writing—review & editing. Xingjie Hao: Conceptualization, Project administration, Writing—review & editing, Supervision. Chen Chen: Conceptualization, Methodology, Project administration, Writing—review & editing, Supervision. Qian Liu: Funding acquisition, Project administration, Writing—review & editing, Supervision.

## References

1. Tieu BC, Lee C, Sun H, LeJeune W, Recinos A, Ju X, Spratt H, Guo D-C, Milewicz D, Tilton RG, et al. An adventitial IL-6/MCP1 amplification loop accelerates macrophage-mediated vascular inflammation leading to aortic dissection in mice. J Clin Invest. 2009;119:3637–3651.

2. Wu Q, Cheng Z, Zhou Y, Zhao Y, Li J, Zhou X, Peng H, Zhang G, Liao X, Fu X. A novel STAT3 inhibitor attenuates angiotensin II-induced abdominal aortic aneurysm progression in mice through modulating vascular inflammation and autophagy. Cell Death Dis. 2020;11:131.

3. Jia Y, Zhang L, Liu Z, Mao C, Ma Z, Li W, Yu F, Wang Y, Huang Y, Zhang W, et al. Targeting macrophage TFEB-14-3-3 epsilon Interface by naringenin inhibits abdominal aortic aneurysm. Cell Discov. 2022;8:21.

4. Yan H, Cui B, Zhang X, Fu X, Yan J, Wang X, Lv X, Chen Z, Hu Z. Antagonism of toll-like receptor 2 attenuates the formation and progression of abdominal aortic aneurysm. Acta Pharm Sin B. 2015;5:176–187.

5. Baxter BT, Matsumura J, Curci JA, McBride R, Larson L, Blackwelder W, Lam D, Wijesinha M, Terrin M. Effect of Doxycycline on Aneurysm Growth Among Patients With Small Infrarenal Abdominal Aortic Aneurysms. JAMA. 2020;323:2029–2038.

6. He X, Wang S, Li M, Zhong L, Zheng H, Sun Y, Lai Y, Chen X, Wei G, Si X, et al. Long noncoding RNA GAS5 induces abdominal aortic aneurysm formation by promoting smooth muscle apoptosis. Theranostics. 2019;9:5558–5576.

7. Zhang T-T, Lei Q-Q, He J, Guan X, Zhang X, Huang Y, Zhou Z-Y, Fan R-X, Wang T, Li C-X, et al. Bestrophin3 Deficiency in Vascular Smooth Muscle Cells Activates MEKK2/3–MAPK Signaling to Trigger Spontaneous Aortic Dissection. Circulation. 2023;148:589–606.

8. Villahoz S, Yunes-Leites PS, Méndez-Barbero N, Urso K, Bonzon-Kulichenko E, Ortega S, Nistal JF, Vazquez J, Offermanns S, Redondo JM, et al. Conditional deletion of Rcan1 predisposes to hypertension-mediated intramural hematoma and subsequent aneurysm and aortic rupture. Nat Commun. 2018;9:4795.

9. Schwaerzer GK, Kalyanaraman H, Casteel DE, Dalton ND, Gu Y, Lee S, Zhuang S, Wahwah N, Schilling JM, Patel HH, et al. Aortic pathology from protein kinase G activation is prevented by an antioxidant vitamin B12 analog. Nat Commun. 2019;10:3533.

10. Huang K, Wu Y, Zhang Y, Youn JY, Cai H. Combination of folic acid with nifedipine is completely effective in attenuating aortic aneurysm formation as a novel oral medication. Redox Biol. 2022;58:102521.

11. Bollache E, Bargiotas I, Giron A, De Cesare A, Soulat G, Mousseaux E, Kachenoura N. Noninvasive quantification of aortic wave reflection timing indices in aging: a study combining MRI and applanation tonometry. J Hypertens. 2025;43:1658–1665.

12. Zhong P, Zhang C, Wu Q, Chang X. Shared genetic loci connect cardiovascular disease with blood pressure and lipid traits in East Asian populations. Front Genet. 2025;16:1635378.

13. Yang T, Yuan X, Gao W, Lu M-J, Hu M-J, Sun H-S. Causal effect of hypertension and blood pressure on aortic diseases: evidence from Mendelian randomization. Hypertens Res. 2023;46:2203–2212.

14. Golledge J, Singh TP. Effect of blood pressure lowering drugs and antibiotics on abdominal aortic aneurysm growth: a systematic review and meta-analysis. Heart. 2021;107:1465–1471.

15. Ma T, Cai Z, Xu X, Cao L, Wang A, Zhang Z, Zhang S, Huang Z, Luo J, Lin S, et al. Calcium channel blockers increase the risk of aortic aneurysm and dissection. Nat Commun [Internet]. 2025 [cited 2025 Dec 29];Available from: https://www.nature.com/articles/s41467-025-68086-5

16. Ho FK, Mark PB, Lees JS, Pell JP, Strawbridge RJ, Kimenai DM, Mills NL, Woodward M, McMurray JJV, Sattar N, et al. A Proteomics-Based Approach for Prediction of Different Cardiovascular Diseases and Dementia. Circulation. 2025;151:277–287.

17. Zhang X, Zhao H, Wan M, Man J, Zhang T, Yang X, Lu M. Associations of 2923 plasma proteins with incident inflammatory bowel disease in a prospective cohort study and genetic analysis. Nat Commun. 2025;16:2813.

18. Zheng Y, Li J, Li Y, Wang J, Suo C, Jiang Y, Jin L, Xu K, Chen X. Plasma proteomic profiles reveal proteins and three characteristic patterns associated with osteoporosis: A prospective cohort study. Journal of Advanced Research [Internet]. 2024 [cited 2025 June 16];Available from: https://www.sciencedirect.com/science/article/pii/S2090123224004740

19. Li Y, Li D, Lin J, Zhou L, Yang W, Yin X, Xu C, Cao Z, Wang Y. Proteomic signatures of type 2 diabetes predict the incidence of coronary heart disease. Cardiovasc Diabetol. 2025;24:120.

20. Cho Y, Haycock PC, Sanderson E, Gaunt TR, Zheng J, Morris AP, Davey Smith G, Hemani G. Exploiting horizontal pleiotropy to search for causal pathways within a Mendelian randomization framework. Nat Commun. 2020;11:1010.

21. Allen NE, Lacey B, Lawlor DA, Pell JP, Gallacher J, Smeeth L, Elliott P, Matthews PM, Lyons RA, Whetton AD, et al. Prospective study design and data analysis in UK Biobank. Sci Transl Med. 2024;16:eadf4428.

22. Sun BB, Chiou J, Traylor M, Benner C, Hsu Y-H, Richardson TG, Surendran P, Mahajan A, Robins C, Vasquez-Grinnell SG, et al. Plasma proteomic associations with genetics and health in the UK Biobank. Nature. 2023;622:329–338.

23. Hibino M, Otaki Y, Kobeissi E, Pan H, Hibino H, Taddese H, Majeed A, Verma S, Konta T, Yamagata K, et al. Blood Pressure, Hypertension, and the Risk of Aortic Dissection Incidence and Mortality: Results From the J-SCH Study, the UK Biobank Study, and a Meta-Analysis of Cohort Studies. Circulation. 2022;145:633–644.

24. Said MA, Verweij N, van der Harst P. Associations of Combined Genetic and Lifestyle Risks With Incident Cardiovascular Disease and Diabetes in the UK Biobank Study. JAMA Cardiol. 2018;3:693–702.

25. Yin Y, Jiang Y, Lam K-WG, Berletch JB, Disteche CM, Noble WS, Steemers FJ, Camerini-Otero RD, Adey AC, Shendure J. High-throughput single cell sequencing with linear amplification. Mol Cell. 2019;76:676–690.e10.

26. Du Y, Guan Y, Shao Z, Jiang M, Qu M, Kong Y, Wu H, Luo D, Peng S, Li S, et al. Integrative Genome-wide Association Meta-analysis of Aortic Aneurysm and Dissection Identifies Five Novel Genes. Genomics, Proteomics & Bioinformatics. 2025;qzaf039.

27. Kurki MI, Karjalainen J, Palta P, Sipilä TP, Kristiansson K, Donner KM, Reeve MP, Laivuori H, Aavikko M, Kaunisto MA, et al. FinnGen provides genetic insights from a well-phenotyped isolated population. Nature. 2023;613:508–518.

28. Ferkingstad E, Sulem P, Atlason BA, Sveinbjornsson G, Magnusson MI, Styrmisdottir EL, Gunnarsdottir K, Helgason A, Oddsson A, Halldorsson BV, et al. Large-scale integration of the plasma proteome with genetics and disease. Nat Genet. 2021;53:1712–1721.

29. Hemani G, Tilling K, Davey Smith G. Orienting the causal relationship between imprecisely measured traits using GWAS summary data. PLoS Genet. 2017;13:e1007081.

30. Jeon CY, Murray MB. Diabetes Mellitus Increases the Risk of Active Tuberculosis: A Systematic Review of 13 Observational Studies. PLoS Med. 2008;5:e152.

31. Burgess S, Thompson SG. Interpreting findings from Mendelian randomization using the MR-Egger method. Eur J Epidemiol. 2017;32:377–389.

32. Cai H, Li H, Xiao X, Wang S, Liu R, Qin Y, Zhou Y, Yao C. TRAF6 promotes abdominal aortic aneurysm development by activating macrophage pyroptosis via the NLRP3/Caspase1/GSDMD pathway. FASEB J. 2025;39:e70318.

33. Butler A, Hoffman P, Smibert P, Papalexi E, Satija R. Integrating single-cell transcriptomic data across different conditions, technologies, and species. Nat Biotechnol. 2018;36:411–420.

34. Ke G, Meng Q, Finely T, Wang T, Chen W, Ma W, Ye Q, Liu T-Y. LightGBM: A Highly Efficient Gradient Boosting Decision Tree [Internet]. 2017 [cited 2025 Nov 12]. Available from: https://www.microsoft.com/en-us/research/publication/lightgbm-a-highly-efficient-gradient-boosting-decision-tree/

35. Lundberg S, Lee S-I. A Unified Approach to Interpreting Model Predictions [Internet]. arXiv.org. 2017 [cited 2025 Nov 12];Available from: https://arxiv.org/abs/1705.07874v2

36. Li M, He X, Gong W, Xiao S, Fu K, Qin Q, Wang L, Li X, Shu C, Li J, et al. Plasma proteomics profiles predict the risk of future aortic aneurysm and aortic dissection. Int J Surg. 2025;

37. Szklarczyk D, Gable AL, Nastou KC, Lyon D, Kirsch R, Pyysalo S, Doncheva NT, Legeay M, Fang T, Bork P, et al. The STRING database in 2021: customizable protein-protein networks, and functional characterization of user-uploaded gene/measurement sets. Nucleic Acids Res. 2021;49:D605–D612.

38. Wu T, Hu E, Chen M, Guo P, Dai Z, Feng T, Zhou L, Tang W, Zhan L, Fu X, et al. clusterProfiler 4.0: A universal enrichment tool for interpreting omics data. Innovation (Camb). 2021;2:100141.

39. Cannon M, Stevenson J, Stahl K, Basu R, Coffman A, Kiwala S, McMichael JF, Kuzma K, Morrissey D, Cotto K, et al. DGIdb 5.0: rebuilding the drug-gene interaction database for precision medicine and drug discovery platforms. Nucleic Acids Res. 2024;52:D1227–D1235.

40. Lu Y, Li Z, Zhang S, Zhang T, Liu Y, Zhang L. Cellular mitophagy: Mechanism, roles in diseases and small molecule pharmacological regulation. Theranostics. 2023;13:736–766.

41. Colman L, Caggiani M, Leyva A, Bresque M, Liechocki S, Maya-Monteiro CM, Mazal D, Batthyany C, Calliari A, Contreras P, et al. The protein Deleted in Breast Cancer-1 (DBC1) regulates vascular response and formation of aortic dissection during Angiotensin II infusion. Sci Rep. 2020;10:6772.

42. Simard E, Söllradl T, Maltais J-S, Boucher J, D’Orléans-Juste P, Grandbois M. Receptor for Advanced Glycation End-Products Signaling Interferes with the Vascular Smooth Muscle Cell Contractile Phenotype and Function. PLoS One. 2015;10:e0128881.

43. Lin B, Ser HL, Wang L, Li J, Chan K-G, Lee L-H, Tan LT-H. The Emerging Role of MMP12 in the Oral Environment. Int J Mol Sci. 2023;24:4648.

44. Liu C, Zhang C, Jia L, Chen B, Liu L, Sun J, Zhang W, You B, Li Y, Li P, et al. Interleukin-3 stimulates matrix metalloproteinase 12 production from macrophages promoting thoracic aortic aneurysm/dissection. Clin Sci (Lond). 2018;132:655–668.

45. Proietta M, Tritapepe L, Cifani N, Ferri L, Taurino M, Del Porto F. MMP-12 as a new marker of Stanford-A acute aortic dissection. Ann Med. 2014;46:44–48.

46. Qi Y, Jiang H, Lun Y, Gang Q, Shen S, Zhang H, Liu M, Wang Y, Zhang J. Protein Drug Targets for Abdominal Aortic Aneurysm and Proteomic Associations Between Modifiable Risk Factors and Abdominal Aortic Aneurysm. J Am Heart Assoc. 2025;14:e037802.

47. Tomaszewski M, Charchar FJ, Nelson CP, Barnes T, Denniff M, Kaiser M, Debiec R, Christofidou P, Rafelt S, van der Harst P, et al. Pathway analysis shows association between FGFBP1 and hypertension. J Am Soc Nephrol. 2011;22:947–955.

48. Tassi E, Lai EY, Li L, Solis G, Chen Y, Kietzman WE, Ray PE, Riegel AT, Welch WJ, Wilcox CS, et al. Blood Pressure Control by a Secreted FGFBP1 (Fibroblast Growth Factor-Binding Protein). Hypertension. 2018;71:160–167.

49. Hiraoka N, Yamazaki-Itoh R, Ino Y, Mizuguchi Y, Yamada T, Hirohashi S, Kanai Y. CXCL17 and ICAM2 Are Associated With a Potential Anti-Tumor Immune Response in Early Intraepithelial Stages of Human Pancreatic Carcinogenesis. Gastroenterology. 2011;140:310–321.e4.

50. Li L, Yan J, Xu J, Liu C-Q, Zhen Z-J, Chen H-W, Ji Y, Wu Z-P, Hu J-Y, Zheng L, et al. CXCL17 expression predicts poor prognosis and correlates with adverse immune infiltration in hepatocellular carcinoma. PLoS One. 2014;9:e110064.

51. Burkhardt AM, Tai KP, Flores-Guiterrez JP, Vilches-Cisneros N, Kamdar K, Barbosa-Quintana O, Valle-Rios R, Hevezi PA, Zuñiga J, Selman M, et al. CXCL17 is a mucosal chemokine elevated in idiopathic pulmonary fibrosis that exhibits broad antimicrobial activity. J Immunol. 2012;188:6399–6406.

52. Oka T, Sugaya M, Takahashi N, Takahashi T, Shibata S, Miyagaki T, Asano Y, Sato S. CXCL17 Attenuates Imiquimod-Induced Psoriasis-like Skin Inflammation by Recruiting Myeloid-Derived Suppressor Cells and Regulatory T Cells. J Immunol. 2017;198:3897–3908.

53. Cao R, Qi W, Huang X, Zheng Y, Zheng R, Ma Y, Zhang H. The Dual Role of the TRAIL-DR5 Signaling Axis in Cardiovascular Disease: From Molecular Mechanisms to Targeted Therapies. Biologics. 2025;19:613–629.

54. Ay H, Arsava EM, Saribaş O. Creatine kinase-MB elevation after stroke is not cardiac in origin: comparison with troponin T levels. Stroke. 2002;33:286–289.

55. Morentin Campillo B, Molina Aguilar P, Monzó Blasco A, Laborda Gálvez JL, Arrieta Pérez J, Sancho Jiménez J, Lamas Ruiz J, Lucena Romero J. Sudden Death Due to Thoracic Aortic Dissection in Young People: A Multicenter Forensic Study. Rev Esp Cardiol (Engl Ed). 2019;72:553–561.

56. Melenovsky V, Adamira M, Kautznerova D, Voska L, Weichet J, Loeys B, Pirk J. Aortic dissection in a young man with Loeys-Dietz syndrome. J Thorac Cardiovasc Surg. 2008;135:1174–1175, 1175.e1.

57. Ono M, Goerler H, Boethig D, Westhoff-Bleck M, Breymann T. Current surgical management of ascending aortic aneurysm in children and young adults. Ann Thorac Surg. 2009;88:1527–1533.

58. Paraskevas KI. Abdominal aortic aneurysms in women. Lancet. 2017;390:1643.

59. Odenbach J, Wang X, Cooper S, Chow FL, Oka T, Lopaschuk G, Kassiri Z, Fernandez-Patron C. MMP-2 mediates angiotensin II-induced hypertension under the transcriptional control of MMP-7 and TACE. Hypertension. 2011;57:123–130.

60. Babey ME, Krause WC, Chen K, Herber CB, Torok Z, Nikkanen J, Rodriguez R, Zhang X, Castro-Navarro F, Wang Y, et al. A maternal brain hormone that builds bone. Nature. 2024;632:357–365.

61. Perbal B. CCN proteins: A centralized communication network. J Cell Commun Signal. 2013;7:169–177.

62. Zhang C, van der Voort D, Shi H, Zhang R, Qing Y, Hiraoka S, Takemoto M, Yokote K, Moxon JV, Norman P, et al. Matricellular protein CCN3 mitigates abdominal aortic aneurysm. J Clin Invest. 2016;126:1282–1299.

63. Pan T-C, Zhang R-Z, Markova D, Arita M, Zhang Y, Bogdanovich S, Khurana TS, Bönnemann CG, Birk DE, Chu M-L. COL6A3 protein deficiency in mice leads to muscle and tendon defects similar to human collagen VI congenital muscular dystrophy. J Biol Chem. 2013;288:14320–14331.

64. Yoshiji S, Lu T, Butler-Laporte G, Carrasco-Zanini-Sanchez J, Su C-Y, Chen Y, Liang K, Willett JDS, Wang S, Adra D, et al. Integrative proteogenomic analysis identifies COL6A3-derived endotrophin as a mediator of the effect of obesity on coronary artery disease. Nat Genet. 2025;57:345–357.

65. Priyanka K, Singh S, Gill K. Paraoxonase 3: Structure and Its Role in Pathophysiology of Coronary Artery Disease. Biomolecules. 2019;9:817.

66. Schweikert E-M, Devarajan A, Witte I, Wilgenbus P, Amort J, Förstermann U, Shabazian A, Grijalva V, Shih DM, Farias-Eisner R, et al. PON3 is upregulated in cancer tissues and protects against mitochondrial superoxide-mediated cell death. Cell Death Differ. 2012;19:1549–1560.

67. Choleva M, Argyrou C, Detopoulou M, Donta M-E, Gerogianni A, Moustou E, Papaemmanouil A, Skitsa C, Kolovou G, Kalogeropoulos P, et al. Effect of Moderate Wine Consumption on Oxidative Stress Markers in Coronary Heart Disease Patients. Nutrients. 2022;14:1377.

68. Qu H-L, Hasen G-W, Hou Y-Y, Zhang C-X. THBS2 promotes cell migration and invasion in colorectal cancer via modulating Wnt/β-catenin signaling pathway. Kaohsiung J Med Sci. 2022;38:469–478.

69. Ishikawa T, Kramer RH. Sdc1 negatively modulates carcinoma cell motility and invasion. Exp Cell Res. 2010;316:951–965.

70. Song Y, Xie Y, Liu F, Zhao C, Yu R, Ban S, Ye Q, Wen J, Wan H, Li X, et al. Expression of matrix metalloproteinase-12 in aortic dissection. BMC Cardiovasc Disord. 2013;13:34.

71. Weis-Müller BT, Modlich O, Drobinskaya I, Unay D, Huber R, Bojar H, Schipke JD, Feindt P, Gams E, Müller W, et al. Gene expression in acute Stanford type A dissection: a comparative microarray study. J Transl Med. 2006;4:29.

72. Kim JH, Kim H, Kim YH, Chung W-S, Suh JK, Kim SJ. Antioxidant Effect of Captopril and Enalapril on Reactive Oxygen Species-Induced Endothelial Dysfunction in the Rabbit Abdominal Aorta. Korean J Thorac Cardiovasc Surg. 2013;46:14–21.

73. Shu C, Zheng X, Wang Y, Xu Y, Zhang D, Deng S. Captopril inhibits matrix metalloproteinase activity and improves dentin bonding durability. Clin Oral Invest. 2022;26:3213–3225.

74. Wang Y, Fu X, Xu J, Wang Q, Kuang H. Systems pharmacology to investigate the interaction of berberine and other drugs in treating polycystic ovary syndrome. Sci Rep. 2016;6:28089.

75. Qiu S, Liu Z, Jiang W-D, Sun J-H, Liu Z-Q, Sun X-D, Wang C-T, Liu W. Diabetes and aortic dissection: unraveling the role of 3-hydroxybutyrate through mendelian randomization. Cardiovasc Diabetol. 2024;23:159.

76. Nienaber CA. Diabetes mellitus and thoracic aortic disease: are people with diabetes mellitus protected from acute aortic dissection? J Am Heart Assoc. 2012;1:e001404.

77. Hibino M, Nienaber CA. Hypertension and diabetes versus the risk of aortic disease: a new look on prevention? Eur J Prev Cardiol. 2022;29:2336–2337.

78. Lindeman JHN, Abdul-Hussien H, van Bockel JH, Wolterbeek R, Kleemann R. Clinical trial of doxycycline for matrix metalloproteinase-9 inhibition in patients with an abdominal aneurysm: doxycycline selectively depletes aortic wall neutrophils and cytotoxic T cells. Circulation. 2009;119:2209–2216.

79. Petrinec D, Liao S, Holmes DR, Reilly JM, Parks WC, Thompson RW. Doxycycline inhibition of aneurysmal degeneration in an elastase-induced rat model of abdominal aortic aneurysm: preservation of aortic elastin associated with suppressed production of 92 kD gelatinase. J Vasc Surg. 1996;23:336–346.

